# Clinical practice guidelines and recommendations in the context of the COVID-19 pandemic: systematic review and critical appraisal

**DOI:** 10.1101/2020.06.19.20134767

**Authors:** Tanja A Stamm, Margaret R Andrews, Erika Mosor, Valentin Ritschl, Linda C Li, Jasmin K Ma, Adalberto Campo Arias, Sarah Baker, Nicola W Burton, Mohammad Eghbali, Natalia Fernandez, Ricardo Ferreira, Gabriele Gäbler, Souzi Makri, Sandra Mintz, Rikke Moe, Elizabeth Morasso, Susan L Murphy, Simiso Ntuli, Maisa Omara, Miguel Angel Simancas Pallares, Jen Horonieff, Gerald Gartlehner

## Abstract

**Background:** The number of published clinical practice guidelines and recommendations related to SARS-CoV-2 infections causing COVID-19 has rapidly increased. However, insufficient consideration of appropriate methodologies in the guideline development could lead to misleading information, uncertainty among professionals, and potentially harmful actions for patients.

**Purpose:** Rapid systematic review of clinical practice guidelines and recommendations in the context of COVID-19 to explore if basic methodological standards of guideline development have been met.

**Data sources:** MEDLINE [PubMed], CINAHL [Ebsco], Trip and manual search; from Feb 1^st^ 2020 until April 27^th^ 2020.

**Study selection:** All types of healthcare workers providing any kind of healthcare to any patient population in any setting.

**Data extraction:** At least two reviewers independently extracted guideline characteristics, conducted critical appraisal according to *The Appraisal of Guidelines for Research and Evaluation Instrument* (AGREE II) and classified the guidelines using the *Association of the Scientific Medical Societies (AWMF) Guidance Manual and Rules for Guideline Development*. We plan six-month updates (living review).

**Data synthesis:** There were 1342 titles screened and 188 guidelines included. The highest average AGREE II domain score was 89% for *scope and purpose*, the lowest for *rigor of development* (25%). Only eight guidelines (4%) were based on a systematic literature search and a structured consensus process by representative experts (classified as the highest methodological quality, S3 according to AWMF). Patients were only included in the development of one guideline. A process for regular updates was described in 27 guidelines (14%).

**Limitations:** Methodological focus only.

**Conclusions:** Despite clear scope, most publications fell short of basic methodological standards of guideline development. Future research should monitor the evolving methodological quality of the guidelines and their updates over time.

**Registration/Publication:** The protocol was published at www.researchgate.net, DOI: 10.13140/RG.2.2.21293.51689. Preliminary results are publicly available on medRxiv.

## Introduction

The novel Coronavirus Disease 2019 (COVID-19) spread rapidly worldwide and the number of cases increased globally at an accelerated rate (1, 2). While measures are essential to mitigate the impact of the pandemic specifically, we also need immediate and targeted action to continue effective and safe healthcare generally. Necessary actions to avoid collateral damages to patients range from adapting healthcare to maximizing safety while providing continuation of usual healthcare to rapidly restart routine care in the areas where healthcare has been reduced due to mandated lockdowns. Moreover, healthcare workers in close physical contact with patients could be infected or could be unknowingly carriers of the SARS-CoV-2 virus themselves.

Clinical practice guidelines and recommendations are needed to support therapeutic decisions and provide an essential knowledge source, especially in such a new and challenging situation. Clinical practice guidelines are statements that include recommendations intended to optimize patient care which are informed by a systematic review of evidence and an assessment of the benefits and harms of alternative care options (3).

The number of published guidelines and recommendations related to COVID-19 has rapidly increased since February 2020. However, insufficient consideration of appropriate methodologies and rigorous strategies in the guideline development process could lead to misleading information, uncertainty among professionals, and potentially harmful actions for patients (4, 5). The aim of our study was therefore to systematically review and critically appraise clinical practice guidelines and recommendations related to SARS-CoV-2 infections and the delivery of healthcare in the current context of COVID-19 from a methodological perspective.

## Methods

### Data Sources and Searches

We performed a rapid systematic review informed by the Cochrane Rapid Reviews Interim Guidance from the Cochrane Rapid Reviews Methods Group (6) in collaboration with Cochrane Austria, located at the Danube University Krems. We will provide regular six-month updates of our review during the COVID-19 situation and six months thereafter (living review), given the necessity for a short-term action, the currently increasing evidence and the rising number of published guidelines and recommendations in the context of COVID-19. An international expert task force including representatives from each of the six WHO regions (Europe, America, Africa, Eastern-Mediterranean, South-East Asia, Western Pacific) defined research questions, keywords, Medical Subject Headings (MeSH) terms and a search strategy for the rapid systematic review. We searched the MEDical Literature Analysis and Retrieval System OnLINE (MEDLINE) [PubMed], the Cumulative Index to Nursing & Allied Health Literature (CINAHL) [Ebsco] and the Turning Research Into Practice (Trip) database on April 27^th^, 2020 (calendar week 17) for medical guidelines and recommendations in the context of the COVID-19 pandemic published after February 1^st^, 2020. An initial limited search was conducted to verify our selection of search terms and proposed strategy. The results of the initial pilot search were discussed with the task force. The search strategy (Supplement Table A) was adapted accordingly and approved before conducting the literature search in MEDLINE, CINAHL and Trip. To identify grey literature (7), the task force members were asked to indicate any additional guidelines and recommendations not identified in the database queries. In addition, we examined the websites of cross-regional associations and institutions at continent level. The study protocol is publicly available at www.researchgate.net (DOI: 10.13140/RG.2.2.21293.51689). All task force members completed a conflict of interest form.

### Study selection

Inclusion criteria were (i) guidelines and recommendations related to the COVID-19 situation, (ii) all types of healthcare workers providing any kind of healthcare to any patient population, (iii) any setting (acute care, rehabilitation, long-term care, practice, home visits, etc.). We excluded documents that only discuss (i) infection control and exposure safety information, (ii) experimental pharmaceutical treatments, (iii) clinical research recommendations, (iv) ethical guidelines, (v) results not produced in direct relation to the COVID-19 situation, (vi) textbooks, theoretical exercises, case studies, experience write-ups, and (vii) documents not available in English.

Results from the searches in MEDLINE and CINAHL were downloaded in Covidence (www.covidence.org) and duplicates removed thereafter. One reviewer (MA) screened titles and abstracts for basic inclusion and exclusion criteria as described above. To ensure accuracy of the selection process, 60 percent of the titles and abstracts were independently screened by other reviewers (EMos, VR, TS). Disagreements were resolved through consensus. Reasons for exclusion are provided in a Preferred Reporting Items for Systematic Reviews and Meta-Analyses (PRISMA) flowchart (8) in Figure 1 and in more detail in the supplementary material (Supplement Table B). Following a training session, full text screening was carried out by at least two reviewers per document (MA, LL, EMos, JM, MO, VR, TS). For the planned updates (living review), we will use automated text mining (tm) based on the tm package in R (www.r-project.org) to support abstract screening and study selection.

**Figure 1.**
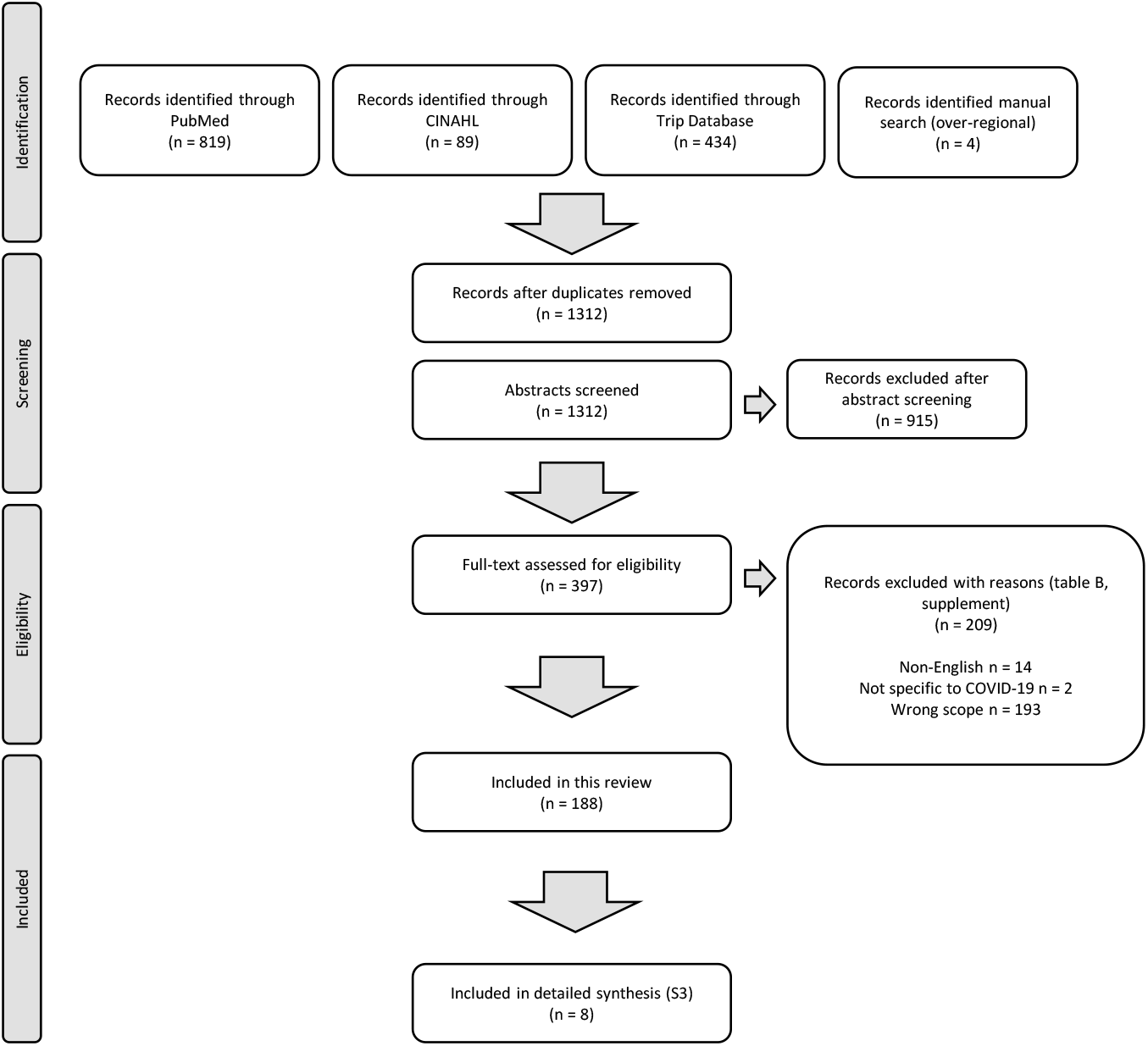
PRISMA chart.

### Data Extraction and Quality Assessment

Data extracted included study characteristics (Supplement Table C), a critical appraisal according to *The Appraisal of Guidelines for Research & Evaluation Instrument* (AGREE II) (4) and classification of the guidelines according to the *Association of the Scientific Medical Societies (AWMF) Guidance Manual and Rules for Guideline Development* (9). The AGREE II is the gold-standard protocol for clinical guideline assessment, development, and reporting (4). It comprises 23 items assessing six domains including scope and purpose, stakeholder involvement, rigor of development, clarity of presentation, applicability, and editorial independence. The AWMF Guidance Manual and Rules for Guideline Development classifies guidelines as being based either on (i) a systematic literature review including a subsequent synthesis of the evidence and a structured consensus process completed by a representative committee (S3), (ii) a systematic literature review and synthesis of the evidence only (S2e), (iii) a structured consensus process completed by a representative committee only (S2k) or (iv) an informal consensus process by a group of experts (S1)(9).

Reviewers pilot-tested the data extraction form on 19 (10%) of the records (Supplement Table C) then independently extracted the data. AGREE II scores were ranked on a Likert scale from 1=strongly disagree to 7=strongly agree. Mean scores between the two reviewers were calculated for each AGREE II item. AGREE II items were summarized into the six domain scores (4). An overall rating and a qualitative recommendation were assigned and disagreements were resolved through consensus. AGREE domain-level scores were calculated for each publication and mean domain-level scores were formed for the entire dataset and the S3 guidelines separately.

### Data Synthesis and Analysis

Descriptive statistics were calculated for relevant extraction fields for the entire dataset using Microsoft Excel, including geographic affiliation of authors, publication status, type of recommendation, target population, focus of recommendation, disease/condition, and setting. Other fields are presented narratively, where appropriate. The world map was created in the Free and Open Source Geographic Information System QGIS (https://qgis.org/en/site/). We used the PRISMA checklist for standardized reporting in systematic reviews and meta-analyses as a reference for the reporting of our results (Supplement Table F).

## Results

In total, we identified 819 records in MEDLINE, 89 in CINAHL and 434 in Trip through our systematic search, and four additional records through other sources (Figure 1). We retained 397 documents for abstract and full text screening. Of these, 188 guidelines and recommendations met the inclusion criteria and were selected for data extraction and critical appraisal.

### Target areas, professionals and country representation of authors

The most frequent medical areas addressed in the guidelines were acute COVID-19 care (n=46; 24%), surgery (n=41; 22%), oncology (n=20; 11%), radiology (n=14; 7%) and cardiology (n=10; 5%). A third (n=57; 30%) of the guidelines focused on miscellaneous other medical areas (Supplement Table D). All except four guidelines (184; 98%) targeted physicians. The four guidelines not targeting physicians referred to care delivered by non-physician health professionals, namely pharmacy practice management for hematopoietic cell transplantation and cellular therapy (10), physiotherapy management for COVID-19 in the acute hospital settings (11) and two articles on pharmaceutical care (12, 13). Overall, 84 guidelines (45%) mentioned the work of non-physician health professionals. This includes naming specific professions and/or grouping all non-physician health professionals under members of a healthcare team. Thirty-two (17%) guidelines also targeted patients and/or the general public. Patients were included as reviewers in the development of only one guideline (14). Experts from 54 countries participated in the guidelines development, with Africa being the least represented continent and North America being the most represented continent (Figure 2). A process for regular updates was described in 27 guidelines (14%).

**Figure 2.**
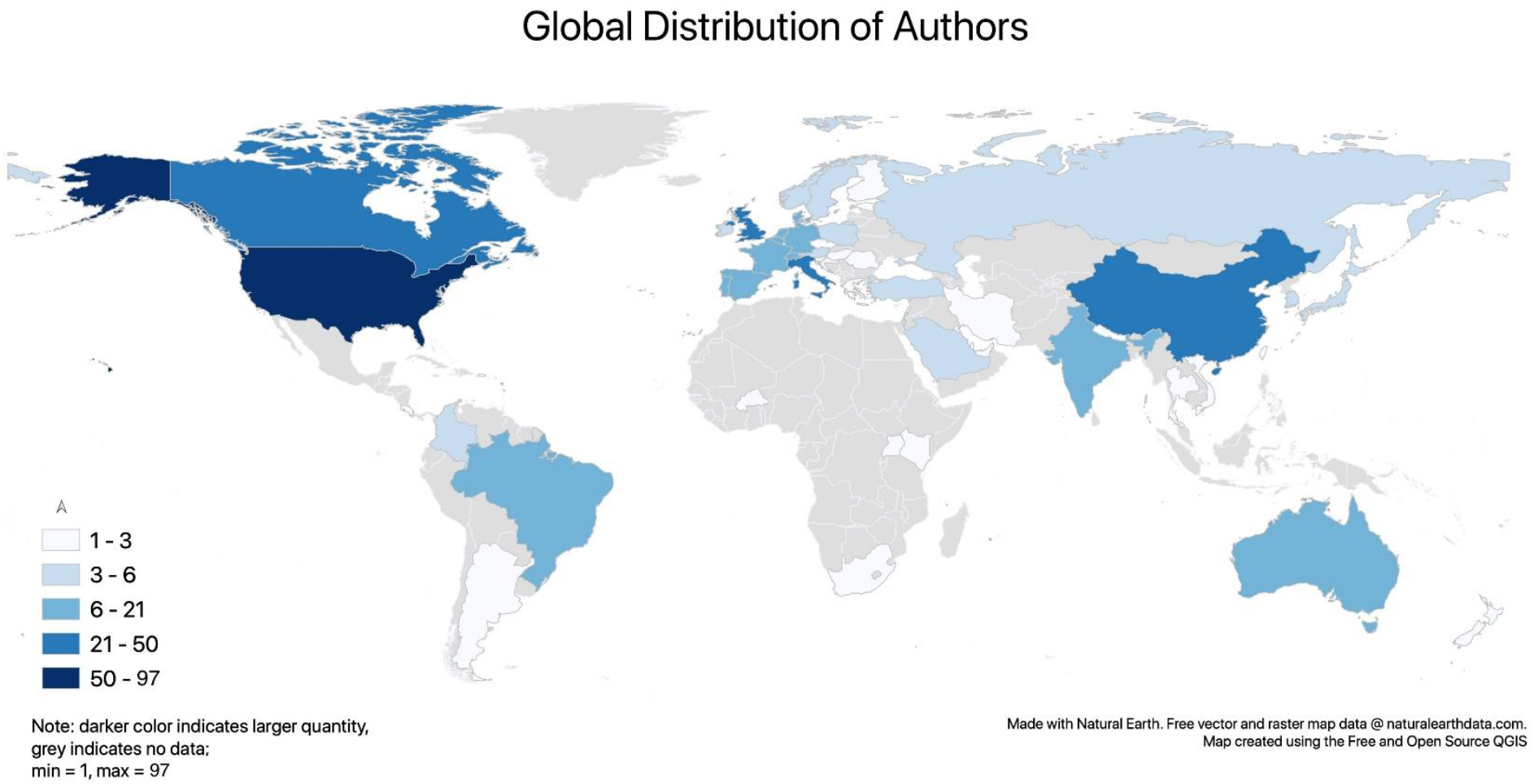
Country representation of the guideline authors. Absolute and relative frequencies per country are shown in supplement Table G.

### Critical methodological appraisal

The highest average AGREE II (4) domain score was given for *scope and purpose* (89%), followed by *clarity of presentation* (70%) and *editorial independence* (61%). *Stakeholder involvement* was on average fulfilled by 43% and *applicability* by 35%. The lowest average AGREE domain score (25%) was given for *rigor of development* due to the fact that methods of development were insufficiently or not reported. The domain scores of each guideline are shown in Supplement Table E. The mean overall quality rating of the guidelines was 3.8 (standard deviation +/-1.4) of a maximum score 7.

Eight of all 188 guidelines (4%) (11, 14–20) were classified as having the highest methodological level (systematic literature search and structured consensus process by representative experts; S3) (9). Twelve guidelines (6%) were solely based on a systematic literature review and synthesis of the evidence (S2e), while a further 12 (6%) were developed using a structured consensus process completed by a representative committee only (S2k). The majority of the guidelines and recommendations, namely 156 (83%), were developed based on an informal consensus process by a group of experts (S1). Papers which referred to a consensus process or expert opinion without providing any details were also assigned to the group of S1 guidelines. An overview about the number of published guidelines regarding their S-classification is depicted separately for each week in Figure 3. The eight S3 guidelines had an average overall quality rating of 6.7 (standard deviation +/-0.5) of a maximum score 7 and are depicted Table 1.

**Table 1.** Data extraction for the eight publications classified as the highest methodological level (systematic literature search and structured consensus process by representative experts; S3). Conflict of interest was declared in all these S3 guidelines. Option for comments or external review were not given in any of these. AGREE II scores were ranked on a Likert scale from 1=strongly disagree to 7=strongly agree.

| Authors, Reference (ID) | Author Countries | Publish Date | Regular Update | History Shown | Methods | Peer-review | Risk of bias - AGREE II Score | Care Focus | Type Focus | Disease/ Condition, if applicable | Setting |
| --- | --- | --- | --- | --- | --- | --- | --- | --- | --- | --- | --- |
| Jin, et al. (19) (358) | China | 06.02.20 | no | no | Literature review and expert consensus | yes | 6 | COVID-19 | COVID-19 | Pneumonia | Generic |
| Gralnek, et al. (16) (302) | Israel, Italy, Germany, Spain, Belgium, UK, the Netherlands, Poland, Greece, Portugal, France, Denmark, Estonia, Slovenia, Croatia, Switzerland, USA | 18.03.20<br>(Up-date 17.04.2020) | yes | yes | Systemic literature review, followed by a formal consensus process by an international group. | yes | 7 | Routine care | Disease-specific | Gastrointestinal endoscopy | Institution |
| Alhazzani, et al. (15) (76) | Canada, Denmark, Saudi Arabia, USA, Netherlands, China, Italy, UAE, Korea, Australia, UK | 28.03.20 | no | no | Systematic literature review and consensus process | yes | 7 | COVID-19 | COVID-19 | Management of critically ill adults with Coronavirus Disease 2019 | Acute / ICU |
| Thomas, et al. (11) (696) | Australia, Belgium, Canada, UK | 30.03.20 | yes | no | Web search for evidence and expert consensus process | yes | 7 | COVID-19 | COVID-19 | Physiotherapy | Acute / ICU |
| Sultan, et al. (14) (678) | USA | 31.03.20 | yes | no | Rapid review and expert consensus | yes | 7 | COVID-19 | Disease-specific | Gastrointestinal procedures | Generic |
| Motlagh, et al. (17) (518) | Iran | 01.04.20 | yes | yes | Literature search and nominal group technique | yes | 6 | Routine care | Disease-specific | Cancer | Generic |
| Rubin, et al. (18) (613) | USA, Canada, South Korea, UK, France, Japan, Germany, Netherlands, Italy, China | 07.04.20 | no | no | A systematic review to inform the discussion of a 15-person expert panel to address 14 questions, corresponding to 11 decision points within 3 scenarios and 3 additional clinical situations. | unclear | 7 | COVID-19 | COVID-19 | Chest imaging | Generic |
| Zhao, et al. (20) (789) | China | 09.04.20 | yes | no | Literature review and expert consensus | yes | 6.5 | COVID-19 | COVID-19 | Respiratory rehabilitation | Generic |

Note. COVID-19= Coronavirus Disease 2019; ICU=intensive care unit; UAE=United Arab Emirates; UK=United Kingdom; USA=United States of America

**Table 2.** Mean AGREE II domain scores for the eight publications classified as the highest methodological level (systematic literature search and structured consensus process by representative experts; S3). AGREE II scores were ranked on a Likert scale from 1=strongly disagree to 7=strongly agree.

| Authors, Reference (ID) | Domain 1<br>Score: Scope<br>& Purpose | Domain 2<br>Score: Stakeholder<br>Involvement | Domain 3<br>Score: Rigor of<br>Development | Domain 4<br>Score: Clarity of<br>Presentation | Domain 5<br>Score:<br>Applicability | Domain 6<br>Score: Editorial<br>Independence | Rate the overall<br>quality of this<br>guideline. | I would<br>recommend this<br>guideline for use. |
| --- | --- | --- | --- | --- | --- | --- | --- | --- |
| Jin, et al. (19) (358) | 100% | 67% | 77% | 83% | 58% | 100% | 6 | Yes |
| Gralnek, et al. (16) (302) | 100% | 78% | 94% | 100% | 58% | 100% | 7 | Yes |
| Alhazzani, et al. (15) (76) | 100% | 47% | 85% | 83% | 67% | 100% | 7 | Yes |
| Thomas, et al. (11) (696) | 100% | 67% | 69% | 100% | 71% | 100% | 7 | Yes |
| Sultan, et al. (14) (678) | 100% | 89% | 96% | 94% | 63% | 100% | 7 | Yes |
| Motlagh, et al. (17) (518) | 97% | 69% | 49% | 67% | 46% | 83% | 6 | Yes |
| Rubin, et al. (18) (613) | 100% | 67% | 81% | 100% | 67% | 100% | 7 | Yes |
| Zhao, et al. (20) (789) | 100% | 56% | 65% | 67% | 75% | 50% | 6.5 | Yes |

**Figure 3.**
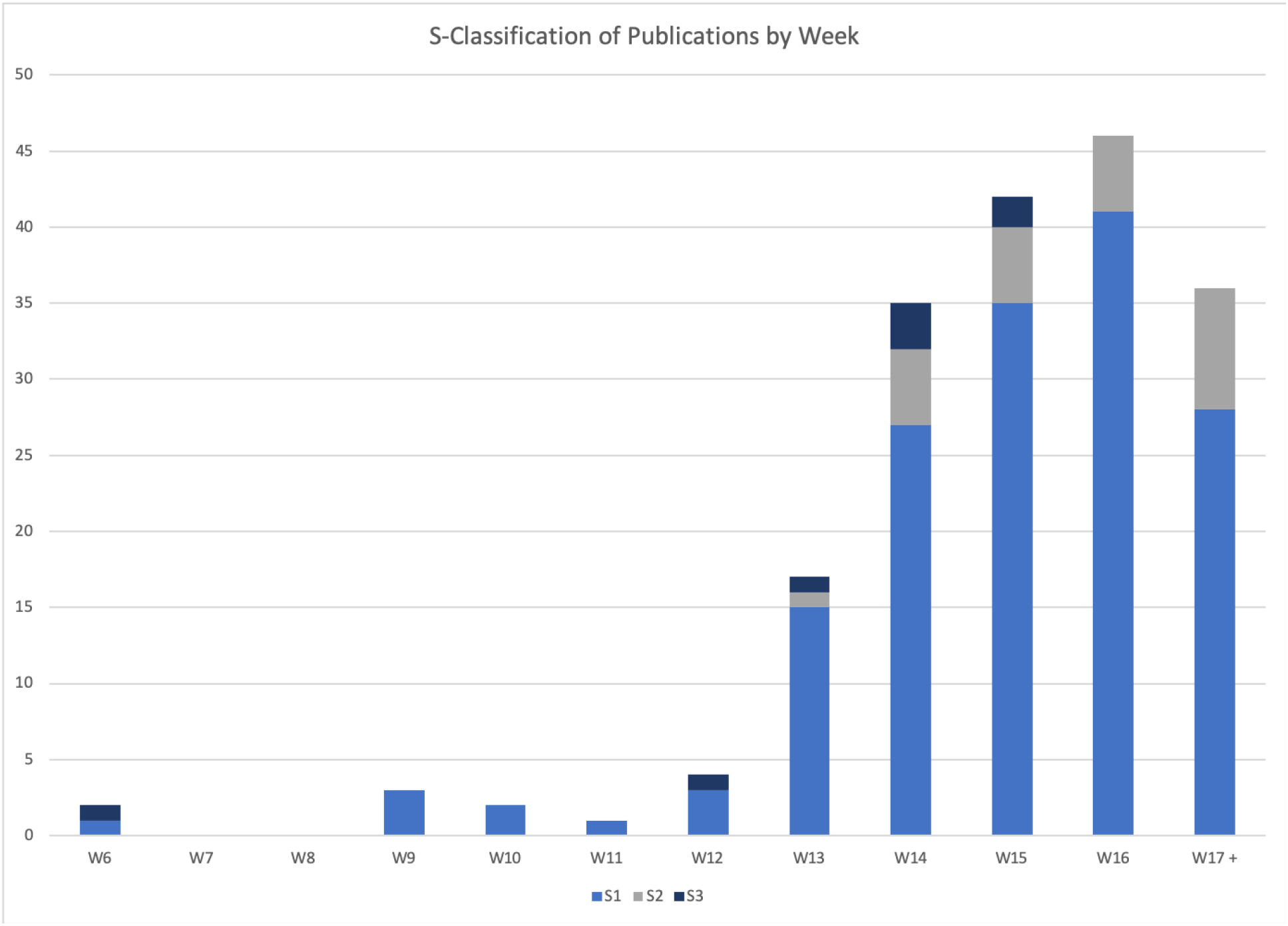
Numbers of published guidelines which increased per week separately for each S-level(9) Guidelines were classified according to the *Association of the Scientific Medical Societies (AWMF) Guidance Manual and Rules for Guideline Development* (9). S3 is a systematic literature review including a subsequent synthesis of the evidence and a structured consensus process completed by a representative committee. S2e refers to a systematic literature review and synthesis of the evidence only; S2k guidelines are based on a structured consensus process completed by a representative committee only. S1 in this work includes both documents based on an informal consensus process by a group of experts as well as expert opinions.

## Discussion

A considerable number of medical guidelines and recommendations related to the COVID-19 pandemic were published from February to April 2020. Despite high AGREE II scores for *scope and purpose* as well as for *clarity of presentation*, the majority of the guidelines and recommendations fell short of basic standards. This was mainly due to a lack of appropriate methodologies and rigorous strategies in the guideline development process. The quality and methodological limitations of the guidelines should be placed in the context of the current unprecedented situation and the human resources required to produce high-quality guidelines in a timely manner. While systematically developed statements reflecting the current state of knowledge and supporting health professionals in their work during the COVID-19 pandemic have been urgently needed, basic methodological standards in the guideline development process are nevertheless essential to avoid misleading information and potentially harmful actions for patients and the healthcare system as a whole. One S3 guideline (19) with high overall methodological ratings had been published in calendar week six from when we found the first guidelines in our search (Figure 3), demonstrating that it was possible to produce high-quality work in a short time. Taking more time to develop guidelines therefore may not always lead to a more rigorous methodological basis. Rather, the methodological commitment, expertise of the authors and engagement of appropriate stakeholders might be important for a rigorous development process, independent of the time period after the outbreak when the guideline was published. Future guideline developers should thus be encouraged to consider rigorous methodological standards; further research could monitor the evolving methodological quality of the guidelines and their updates over time. Similar to our findings of clinical practice guidelines falling short of basic methodological standards, a recent review of prediction models for diagnosis and prognosis of COVID-19 concluded that the published models were also poorly reported and at high risk of bias (21).

Clinical practice guidelines with the highest methodological quality, classified as S3 according to AWMF, are built on both a systemic literature review and a structured consensus process by representative experts. Only 4% of all guidelines in our study were S3. Another 6% reported solely a structured consensus process without a systematic literature review (classified as S2k according to AWMF). The majority (83%) was based on an informal consensus by a group of experts only. However, even if some guidelines developers might argue that well-designed studies were lacking at a certain time point and a systemic literature review as a basis for the guideline development was therefore not conducted, a structured consensus process could still have been done.

A clear updating process, patient and public involvement, a systematic development processes incorporating systematic review, an explicit recommendation development process (rigor of development domain), and guidance on implementation of these guidelines (applicability domain) are needed areas to improve guideline development quality. Given the increasing evidence related to the COVID-19 pandemic, a clear updating process, as well as information of where previous and later versions of the guideline can be found might be essential information which should be explicitly stated in each document. Although patient and public involvement is recognized as a key component of clinical practice guideline development (22), almost no citizens, patients or their public representatives were involved in the development of any of the documents, except for one guideline (14).

Non-physician health professionals provide essential care, especially in the area of non-pharmacological interventions including physical activity/exercise, activity pacing, pain management, complex chronic disease management, nutrition, speech/aural care, personal care, disability, rehabilitation, substance use, aged care and psychosocial health (23). Some of these interventions have been continuously provided during the current COVID-19 situation, some health services have not been delivered due to home quarantine and physical distancing.(24) Guidelines and recommendations on how to manage and respond to this situation are often missing. Currently, the majority of the published guidelines, which we reviewed, did not focus on the specific needs of non-physician health professionals who are usually in direct/face-to-face patient contact. However, due to often unclear information about the background and role of each author in the guideline development process, transparency on stakeholder involvement was sometimes lacking and judgement was left to the discretion of the reviewers. Balanced stakeholder involvement as well as explicit information about this process in the paper would be necessary for a transparent guideline development that takes into account different treatment perspectives.

Our study focused on quality criteria of the published guidelines and recommendations. We did not extract, compare and synthesize the medical content of the recommendations, due to the heterogeneity of medical disciplines included in this work. This might be regarded as a limitation of our work. Nevertheless, our results might also contribute to a further conversation on how best to mobilize national and international resources to develop high-quality guidelines in crisis situations in the future.

## Conclusion

A considerable number of medical guidelines and recommendations were published in the context of the COVID-19 pandemic from February to April 2020. Despite high AGREE II scores for *scope and purpose* as well as for *clarity of presentation*, the majority of the guidelines and recommendations fell short of basic methodological standards. This was mainly due to a lack of appropriate methodologies and rigorous strategies in the guideline development process. Updated guidelines should include up-to-date information, transparent stakeholder involvement, consideration of various patient populations, and rigorous strategies and methodologies.

## Data Availability

All data are published as supplemental files.

## Declaration of interests

We declare no competing interests related to this work.

## Acknowledgement

We would like to thank Mohammed Gamal, Judith Höllebrand, Claudia Oppenauer, Romualdo Ramos, Sinisa Stefanac and Thea Vliet Vlieland for their support in data management and thoughtful advice.

## Details of authors’ contributions

MA, EMos, VR, LL, JM, ME, NF, RF, GGä, SMa, SMi, RMo, SMu, SN, MO, JH, GGa, and TS were responsible for the study concept and design. MA, LL, EMos, JM, MO, VR and TS conducted the literature review and extracted and analysed data. MA, EMos, VR, LL, JM, AA, SB, NB, ME, NF, RF, GGä, SMa, SMi, RMo, EMor, SMu, SN, MO, MSP, JH, GGa, and TS interpreted the data, wrote and reviewed the manuscript. MA, VR and TS were responsible for the tables and figures.

## Details of ethical approval

Not required.

## Patient and public involvement

Three co-authors (JH, SMa and EMor) are patient representatives/advocates and participated in setting up the study concept and design, interpreting the data and writing and reviewing the manuscript.

## Data sharing statement

All relevant data are up-loaded as supplemental files.

## Protocol

The protocol was published at www.researchgate.net, DOI: 10.13140/RG.2.2.21293.51689.

## Funding Source

This study was partly funded by the COVID-19 Rapid Response Funding Scheme of the Wiener Wissenschafts-, Forschungs-und Technologiefonds (WWTF)/Vienna Science and Technology Fund (project number COV20-028). The funding institution had no influence on the results of this work.

## Notes

### Competing Interest Statement

The authors have declared no competing interest.

### Clinical Protocols

https://www.researchgate.net/publication/341670661_Clinical_practice_guidelines_and_recommendations_in_the_context_of_the_COVID-19_pandemic_systematic_review_and_critical_appraisal

